# PATCRdb: Database of TCRs from data mining patent documents

**DOI:** 10.1101/2023.01.05.23284150

**Authors:** Yoona Lee, Rebecca Freitag, Rajkumar Ganesan, Veit Schwämmle, Sandeep Kumar, Konrad Krawczyk

## Abstract

T-cells are crucial actuators of the innate immune system. Because their receptors recognize intracellular disease markers, there is considerable interest in developing them as novel biotherapies. Computational methods to support discovery, design and development of TCR-based therapeutics need robust repositories of curated sequence and structural information on TCRs. The urgency of this need is highlighted by the recent approval of the first TCR biotherapeutic, tebentafusp. In this work, we have collected patent data on TCR sequences to provide early access to TCRs that are in various stages of product and clinical development (pre-FDA approvals) and are already past the initial discovery / proof of concept (scientific publications) stages. We employ literature mining to identify patent documents disclosing TCR sequences. Such documents are further analyzed to provide a birds-eye view of TCR patenting landscape. We compile the information into a database available at http://github.com/konradkrawczyk/patcrdb that we hope should help TCR engineers.

## Introduction

Our immune system is capable of identifying and destroying infected, damaged, as well as malignant cells. Immune cells known as killer T-cells are particularly effective against cancer because of their propensity to bind to antigens on the surface of cancer cells.

This natural action of T-cells can be employed therapeutically via adoptive cell and molecular therapeutics such as CAR-Ts, T-cell engagers, T-cell Receptors, and multi-specific T-cell receptor mimicking antibodies (1). Researchers can improve the ability of a patient’s own T-cells by engineering them to recognize certain molecular markers and destroying tissue where these are detected. This can be achieved by modifying the T-cell receptors (TCRs), that are responsible for molecular recognition abilities of T-cells. TCRs are membrane proteins on T-cell’s outer surfaces that recognize intracellular peptides presented by the major histocompatibility complex (MHC). By engineering TCRs to recognize specific peptide-MHC complexes (pMHC) that are indicative of a disease, the T-cells can be used to destroy malignant cells and tumors.

Despite their potential, some of the early attempts at employing engineered TCRs resulted in extreme toxicity killing patients participating in clinical trials (2,3). Recently, however, the first TCR-based therapy, Immunocore’s tebentafusp, has been approved for uveal melanoma (4). This first success is likely to be followed by several others and development of future T-cell receptor based therapies shall require careful data interpretation to improve the probability of therapeutic success of these molecules and reduce toxicity risks at the same time.

One of the most promising avenues to designing and accelerating the speed of bringing novel TCR-based therapies to the clinic is to employ innovative computational methodologies which hold the promise to engineer TCRs recognition and therapeutic properties without resort to expensive and laborious experimentation (5). Methods are being developed to predict pMHC binding (6), predict the 3D structure (7) or develop TCR-specific large language models (8). Suchy methods provide a theoretical basis for developing TCRs or TCR mimetic antibodies with desired binding properties, without the burden of experimental methods. Development of such methods relies heavily on availability of large and well annotated structural(9) and sequence datasets(10) specific to TCRs. To the best of our knowledge however, there are no databases that specifically contain sequence and structural information on the TCRs and annotate the therapeutic potentials of TCRs. The information on TCR molecules that have the potential to enter clinical development can be found in patents (11) and Investigational New Drug (IND) submissions with the regulatory agencies (4). There are currently seven TCR-based target-recognizing molecules in clinical development (4). Here molecules are characterized by their primary sequence together with the disclosure of their therapeutic action. Thus, patent literature serves as a source to assess the trend of therapeutically relevant TCRs. In this paper, we identify TCR related patents and give an overview of the pre-FDA landscape and build a basis for computational models. We share this information as a database of the sequences and metadata for the benefit of computational TCR engineers, available at http://github.com/konradkrawczyk/patcrdb.

## Materials & Methods

### Collection of raw/original TCR sequences

The TCR sequences from patent documents were extracted using a protocol described previously for antibodies (11). Briefly, well-formatted biological sequence depositions from USPTO, PSIPS, WIPO, and DDBJ were downloaded in January 2022. The amino acid sequences were processed using Hidden Markov Models (12) trained on TCR genes (13), as compiled by ANARCI (14). The nucleotide sequences were processed using IgBLAST (15), also given a reference of TCR genes.

### Quality checks

Both ANARCI and IgBLAST identify TCR sequences by virtue of alignment to germline genes, rejecting other sequences. When a sequence was identified as TCR by either ANARCI or IgBLAST, we checked its quality. We only accept sequences that have all CDRs and do not contain non-standard amino acids, particularly X, which is a common placeholder in patent sequences. Pairing between alpha and beta chains is not possible to infer with the metadata contained in listings and thus all of our sequences are unpaired. Each sequence is represented by the following data transformations:

- **Original sequence**. The sequence in the form ANARCI or IgBLAST received it.
- **Variable region sequence**. After processing the original sequence via ANARCI/IgBLAST, only the variable portion.
- **Sequence name** - name of the patent and the sequence number where the sequence was identified.
- **Families** - European Patent Office (EPO)-defined families, that provide an equivalence class for patent documents with different numbers across different jurisdictions (to be taken as a unique identifier of a patent document).

### Metadata Curation

Whenever a sequence was identified as TCR by IgBLAST or ANARCI and fulfilled our quality criteria, we retrieved the patent document information associated with it through its patent number. Different patent numbers that refer to the same documents are identified by their patent family as defined by the European Patent Office. For each patent family identified in this fashion, we collect the following information:

- **Title**. The title of the patent document.
- **Abstract**. The abstract of the patent document.
- **Family**. The EPO family assigned to the document.
- **Applicants**. The applicants on the patent document, e.g. company names.
- **Inventors**. The people cited as inventors of the given work.
- **Dates**. List of dates associated with the document, as there can be several in a lifetime of a patent document.
- **Classes**. The Cooperative Patent Classification (CPC) scheme grouping.

The TCR sequences and document metadata are linked by their patent family number. Both data abstractions, sequence and metadata, allow to perform analyses of both the sequence space of TCRs in patent documents, as well as an indication of their application by virtue of textual metadata.

## Availability

The PATCRdb is made available under the following github link: http://github.com/konradkrawczyk/patcrdb. The database is provided as a .csv format listing the sequences and their metadata separately. Subsequent updates to the database will be pushed to this repository.

## Results

### Database Content Statistics

We extracted TCR sequences by downloading data from the three patent sources: United States Patent and Trademark Office (USPTO, FT and PSIPS), World Intellectual Property Organization (WIPO), DNA Data Bank of Japan (DDBJ), as described previously (11).

It is common to seek protection under the WIPO Patent Cooperation Treaty (PCT) system to broaden the coverage of patent documents across many jurisdictions around the world. Data from other major jurisdictions such as Japanese and Korean patent offices were included through DDBJ. Notably the EPO and Chinese patents are not included because of unavailability for large-scale free data mining at this stage. Even though EPO and Chinese patents are not included in this database, USPTO and WIPO do cover a large portion of globally relevant patents.

The amino acid sequences were further processed using ANARCI (14) to determine the alpha and beta chain domains. The number of raw sequences quantified, and alpha and beta chains identified are reported in Table 1.

**Table 1.** Published biological sequences and proportions identified as TCR chains. The total number of raw sequences is given in column Total Raw. Full sequences were further divided into alpha and beta domains using ANARCI. All the sequences are unpaired.

| Source | Total Raw | TCR identified<br>(unique alpha($\alpha$), beta($\beta$)) |
| --- | --- | --- |
| USPTO FT | 9,052 | $\alpha = 4,572, \beta = 4,479$ |
| USPTO PSIPS | 3,399 | $\alpha = 1,629, \beta = 1,766$ |
| WIPO | 6,891 | $\alpha = 3,470, \beta = 3,419$ |
| DDBJ | 4,602 | $\alpha = 2,409, \beta = 2,193$ |

TCR sequences from all four sources were combined and filtered to retrieve unique sequences. As a result, 5,023 unique sequences were obtained.

TCR sequences and the patent family that they are identified with, were linked. A patent family is an identification given to a collection of patent applications that have similar technical content. As a result, 5,023 unique TCR sequences were distributed among 495 patent families.

### Patent content analysis

The patent data were investigated by extracting metadata from the patent documents, such as titles, abstracts, inventors, and classifications.

In the first instance the extracted information was utilized to determine the proportion of patents intended for different TCR engineering methods. For instance, a notable way of using T-cells is via Chimeric Antigen Receptors (CAR-T), where an antibody-based receptor is designed attached to the T-cell surface. We manually checked the 495 Titles and Abstracts, whether to our reading the document specifically referred to T-cells (rather than them being used tangentially). Of 495 unique patent families, 306 (62.7 %) were deemed to be T-cell related. Total of 38 (7.7 %) families specifically mentioned CAR-T therapies in the title/abstract. There were 135 (27.2 %) families with TCR and antibody sequences, which in turn gave out 361 (72.8 %) patent families that only contained TCRs and no antibodies.

### Patent application analysis

Cooperative Patent Classification (CPC), an international classification scheme managed by EPO and USPTO in a joint manner, was used to analyze patent classifications. This scheme groups patents using components such as section, class, subclass, main group, and subgroup. For example, classification C07K16/2866 has section C, class 07, subclass K, main group 16, and subgroup 2866. This provides a high-level indication of the purpose of the patent, and thus the molecules contained therein, for instance therapeutic TCRs against certain targets.

CPC designation was extracted from each patent to identify the purpose of each invention. This scheme covered 433 (87.3 %) out of 495 patent families, compiled in Table 2. Subgroup C07K14, which indicates peptides having more than 20 amino acids, was the most common classification, found in 355 (82.0 %) out of 433 patent families. Families listing peptides for medicinal purposes (A61K38) accounted for 192 (44.3 %) families. The more general medicinal classification A61K (preparation for medical purposes) accounted for 336 (77.6 %) of the 433 patent families. This suggests that the majority of patent documents holding TCR sequences are aimed for medical applications.

**Table 2.** Subclasses of the CPC patent classifications. The % in total families is given out of the 433 patent families which were covered by the CPC scheme.

| Class | Total families (%) | Description |
| --- | --- | --- |
| C07K14 | 355 (82.0) | Peptides having more than 20 amino acids; Gastrins; Somatostatins; Melanotropins; Derivatives thereof |
| A61K38 | 192 (44.3) | Medicinal preparations containing peptides |
| A61P35 | 184 (42.5) | Antineoplastic agents |
| A61K35 | 158 (36.5) | Medicinal preparations containing materials or reaction products thereof with undetermined constitution |
| C07K16 | 154 (35.6) | Immunoglobulins [IGs], e.g. monoclonal or polyclonal antibodies |
| C12N5 | 147 (33.9) | Undifferentiated human, animal or plant cells, e.g. cell lines; Tissues; Cultivation or maintenance thereof; Culture media therefor; |
| A61K39 | 138 (31.9) | Medicinal preparations containing antigens or antibodies |
| A61K2039 | 135<br>(31.2) | Medicinal preparations containing antigens or antibodies |
| C07K2319 | 131<br>(30.3) | Fusion polypeptide |
| C12N15 | 129<br>(29.8) | Mutation or genetic engineering; DNA or RNA concerning genetic engineering, vectors, e.g. plasmids, or their isolation, preparation or purification; Use of hosts therefor |
| C07K2317 | 120<br>(27.7) | Immunoglobulins specific features |
| G01N33 | 92<br>(21.2) | Investigating or analysing materials by specific methods not covered by groups |
| C12N2510 | 81<br>(18.7) | Genetically modified cells |
| G01N2333 | 40<br>(9.2) | Assays involving biological materials from specific organisms or of a specific nature |
| C12N2740 | 36<br>(8.3) | Reverse transcribing RNA viruses |
| A61P37 | 36<br>(8.3) | Drugs for immunological or allergic disorders |
| C12Q1 | 31<br>(7.2) | Measuring or testing processes involving enzymes, nucleic acids or microorganisms; Compositions therefor; Processes of preparing such compositions |
| C12N9 | 27<br>(6.2) | Enzymes; Proenzymes; Compositions thereof; Processes for preparing, activating, inhibiting, separating or purifying enzymes |
| A61P31 | 26<br>(6.0) | Antiinfectives, i.e. antibiotics, antiseptics, chemotherapeutics |
| A61K47 | 24<br>(5.5) | Medicinal preparations characterized by the non-active ingredients used, e.g. carriers or inert additives; Targeting or modifying agents chemically bound to the active ingredient |

### Patent inventorship analysis

We identified the patent applicants, such as companies, government organizations that had the most TCR-containing documents in our database. Applicants were sorted into three categories, company, research organization, and government organization. Non-governmental research centers, universities, and hospitals were included under research organization, whereas research centers, projects, or departments under government jurisdiction were classified as government organizations. Patent applications filed by individuals (242 applicants) and inventors that were hard to identify (3 applicants) were removed.

A total of 203 applicants were involved in the TCR patent registration, of which 119 were company inventors (Table 3). The most crucial commercial player was Immunocore, involved with 55 TCR inventions (# patent families), followed by Adaptimmune (44 inventions), Immatics biotechnologies (17 inventions), and Medigene (15 inventions). That is fitting as these companies are on the fore-front of TCR therapies with Immunocore recently being awarded the first approval for TCR-based therapy. Hutchinson Fred cancer research center was the leading applicant in the research sector with 19 inventions filed. University of California came next with 14 inventions involved. Last but not least, of government organizations, the US Department of Health and Human Services showed a notable number of 35 inventions.

**Table 3.** Ranking of top TCR patent applicants. Applicants who have registered the most TCR inventions by the number of patent families in our database.

| **Company applicants** | **#patent families** |
| --- | --- |
| Immunocore | 55 |
| Adaptimmune | 44 |
| Immatics Biotechnologies | 17 |
| Medigene | 15 |
| Hyseq | 12 |
| Guangdong Xiangxue Life Sciences | 11 |
| Innovative Cellular Therapeutics | 10 |
| Altor Bioscience | 9 |
| Nuvelo | 8 |
| Guangzhou Xiangxue Pharmaceutical Co | 7 |
| Tcr2 Therapeutics | 7 |
| Avidex | 6 |
| Res Ass For Biotechnology | 6 |
| Ucl Business | 6 |
| Elstar Therapeutics | 5 |
| Ksq Therapeutics | 5 |
| Gritstone Oncology | 4 |
| Hangzhou Converd Co | 4 |
| Juno Therapeutics | 4 |
| Synimmune | 4 |
| Takara Bio | 4 |
| Ablynx | 3 |
| Adaptive Biotechnologies | 3 |
| Agenus | 3 |
| Biontech Cell & Gene Therapies | 3 |
| Cero Therapeutics | 3 |
| Corixa | 3 |
| Dana Farber Cancer Inst | 3 |
| **Research organization applicants** | **#patent families** |
| Hutchinson Fred Cancer Research Center | 19 |
| Univ California | 14 |
| Max Delbrück Center For Molecular Medicine | 10 |
| Univ Texas | 6 |
| Univ Der Johannes Gutenberg Univ Mainz | 5 |
| Univ Oslo | 4 |
| Univ Berlin Charite | 4 |
| Univ Health Network | 4 |
| Univ Illinois | 4 |
| California Inst Of Technology | 3 |
| Leiden University Medical Center | 3 |
| Tron Translationale Onkologie An Der Univ Der Johannes Gutenberg Univ Mainz Gemeinnuetzige | 3 |
| Univ Basel | 3 |
| Univ Pennsylvania | 3 |
| Univ Washington | 3 |
| Fond Centro San Raffaele | 2 |
| Institute For Systems Biology | 2 |
| La Jolla Institute Allergy & Immunology | 2 |
| San Raffaele Hospital | 2 |
| Univ Columbia | 2 |
| Univ Kyoto | 2 |
| Univ Leland Stanford Junior | 2 |
| Univ Massachusetts | 2 |
| Univ Mie | 2 |
| Univ Minnesota | 2 |
| Univ Notre Dame Du Lac | 2 |
| Univ Southern California | 2 |
| **Government organization applicants** | **#patent families** |
| US Secretary Dept Of Health And Human Service (US) | 35 |
| Guangzhou Institute Biomed & Health (China) | 5 |
| Helix Research Institute (Japan) | 2 |
| National Institute For Health And Medical Research INSERM (France) | 2 |
| Administracion General De La Comunidad Autonoma De Euskadi (Euskadi) | 1 |
| Agency Science Tech & Research (Singapore) | 1 |
| National Center For Scientific Research CNRS (France) | 1 |
| German Cancer Research Center DKFZ (German) | 1 |
| National Cancer Center Japan (Japan) | 1 |
| National Institutes Of Health NIH (US) | 1 |
| Vlaams Instituut Voor Biotechnologie (Belgium) | 1 |

The top four companies were explored for their therapeutic pipelines. Immunocore had many different oncological targets such as uveal and cutaneous melanoma, synovial sarcoma, non-small cell lung cancer (NSCLC), small cell lung cancer (SCLC), breast, endometrial, ovarian, gastric, head and neck, colorectal, and pancreatic cancer. Infectious diseases such as hepatitis B virus (HBV) and human immunodeficiency virus (HIV) were also set as their pipeline. Adaptimmune focused on synovial sarcoma, myxoid/round cell liposarcoma (MRCLS), esophageal and gastroesophageal junction (EGJ) cancer. Immatics and Medigene addressed solid tumors and hematological cancer. All in all, top TCR companies appear to be focusing their efforts on solid tumors, which might be an indication of the upcoming TCR therapies.

### TCR patent documents by year

The year-sorted numbers of TCR-related patent documents were visualized to reveal possible trends in the volume of novel therapies being developed. Since the patent family can be associated with several timestamps we analyzed the earliest submission date and the most recent submission date separately.

Figure 1 left shows patent documents sorted by the earliest submitted year. The earliest documents we could find date to the early 2000s, with a long plateau of limited activity. The number of patent documents containing TCR sequences appears to have started increasing in 2015. Figure 1 b shows the patent documents sorted by their latest date, which appears to continue the trend of more TCR patent families being filed. Altogether this indicates that TCR patent activity increased, and appears to be gaining momentum.

**Figure 1.**
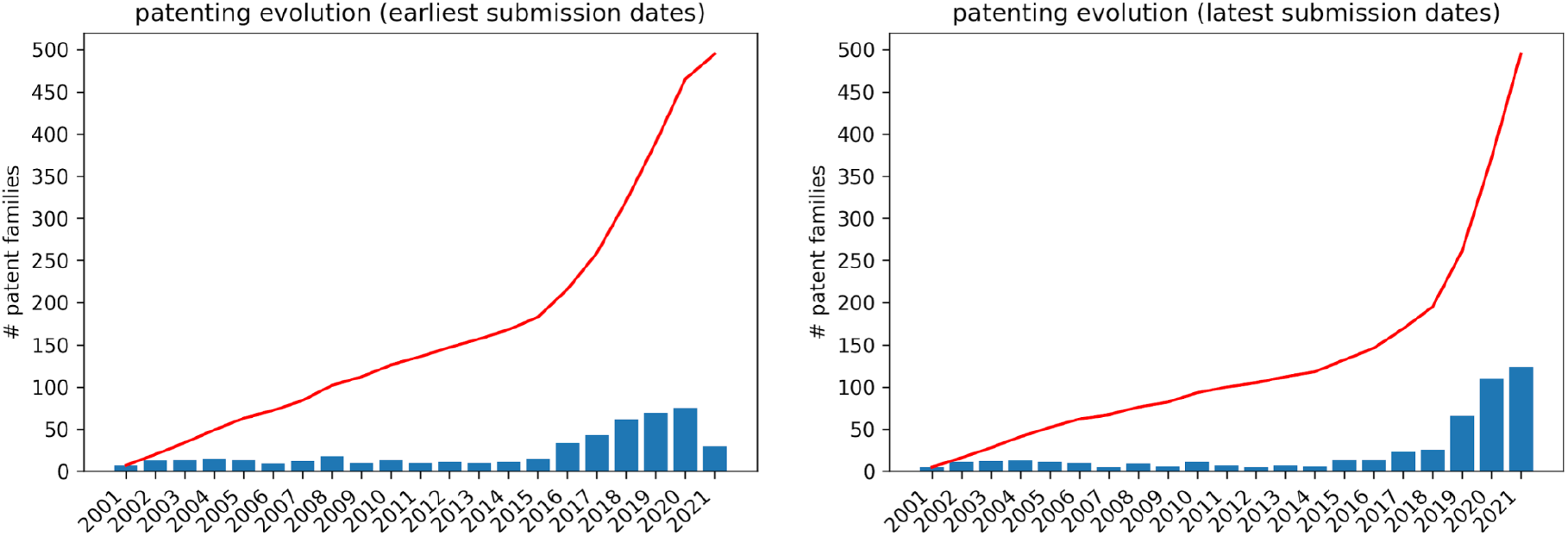
Number of patent submissions in the last 20 years. **Left.** The number of patent families listing TCR sequences by their earliest submission dates. **Right**. The number of patent families listing TCR sequences by their latest submission dates.

### Sequence analysis by gene

The final aspect we considered were the assigned TCR germline genes of the patented sequences. Choice of the germline can be associated with biophysical/therapeutic properties and is seldom left to chance. For this reason, statistics of TCR genes that are the most commonly used in patents could be an indication of the engineering choices taken in designing these molecules for therapeutic purposes.

In Figure 2, we present the most common human TCR V-region genes to which the patent sequences align. The most used human alpha chain V-gene by sequence was TRAV12-2, accounting for 18.25 % of all patented sequences. The most frequently observed human beta chain germline usage was TRBV6-5, accounting for 10.56 % of all patented sequences.

**Figure 2.**
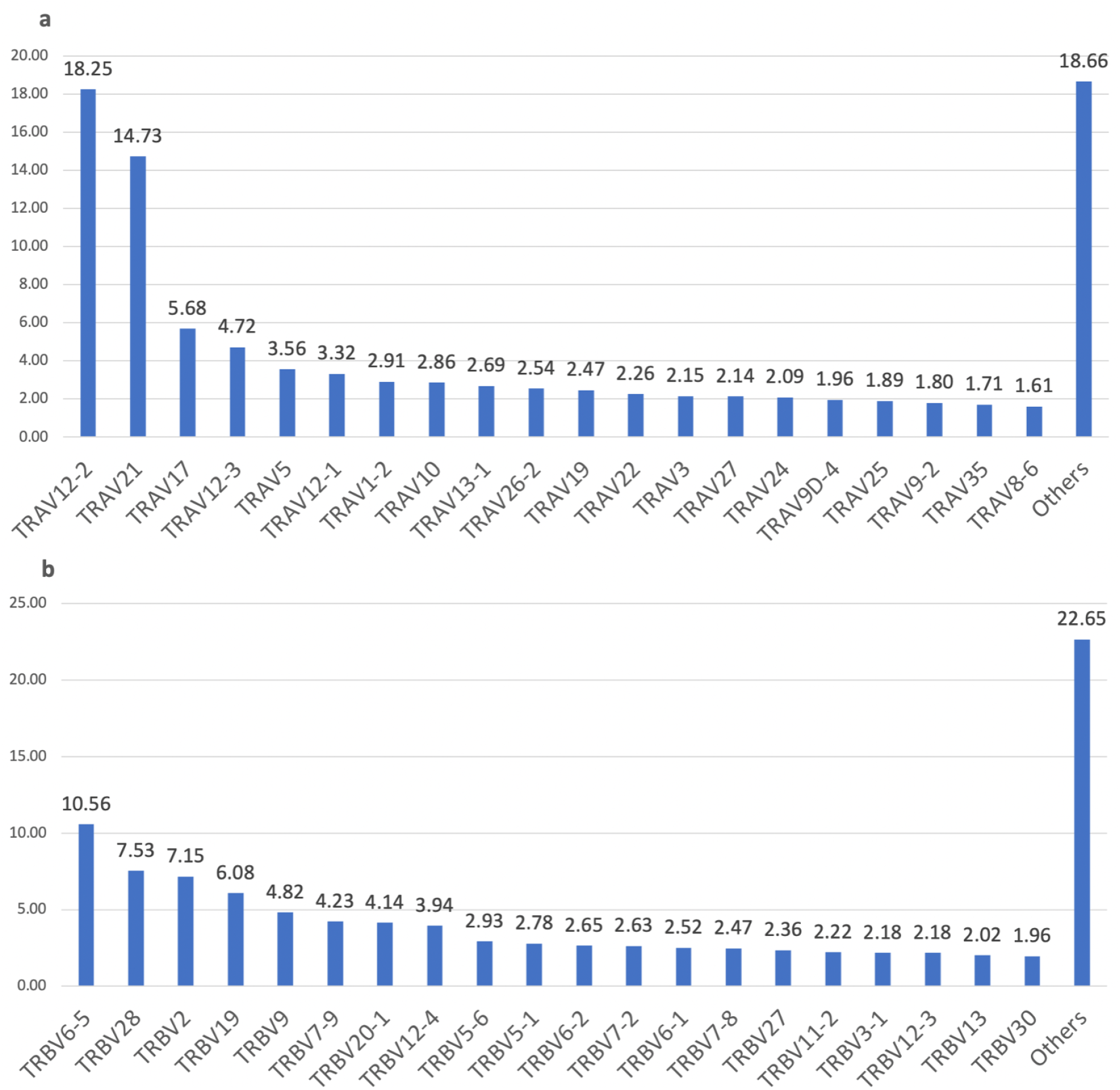
Bar plots of T cell receptor V-gene usage in patented sequences. **a**: TRAV, alpha genes. **b**: TRBV, beta genes.

We contrasted the germline frequency from patents from this observed naturally. Barennes et at. (2021) compared nine T cell receptor repertoire sequencing methods using the blood samples of two healthy male donors (16). As a result, they present the distribution of TRAV and TRBV gene usage. Table 4 shows the ten most used genes for TRAV and TRBV. Comparing their results to Figure 2, we can find 4 TRAV genes, which are part of the most common TCR V-genes in patents. These genes are: TRAV13-1, TRAV21, TRAV12-1, and TRAV17. For TRBV, 8 out of the 10 genes are present in Figure 2 as well: TRBV20-1, TRBV5-1, TRBV7-9, TRBV28, TRBV7-2, TRBV2, TRBV3-1, TRBV6-5.

**Table 4.** Top 10 most common t cell receptor V-region genes as published by Barennes et al. (16)

| TRAV | TRBV |
| --- | --- |
| TRAV13-1 | TRBV20-1 |
| TRAV21 | TRBV29-1 |
| TRAV2 | TRBV5-1 |
| TRAV9-2 | TRBV7-9 |
| TRAV13-2 | TRBV28 |
| TRAV38-2DV8 | TRBV7-2 |
| TRAV29DV5 | TRBV2 |
| TRAV12-1 | TRBV3-1 |
| TRAV17 | TRBV4-1 |
| TRAV26-1 | TRBV6-5 |

In a different study, the TCR-β repertoire of 10 healthy individuals were sequenced, and the following analysis included the usage of the TRBV genes (17). They reported a high variation of TRBV usage frequency for some genes between the individuals, but TRBV20–1, TRBV2, TRBV19, TRBV5–1, and TRBV7–9 tend to show generally higher usage. All of these genes can be found within the first ten most used beta chain V genes in published patents (Figure 2) as well as in Table 4 (except TRBV19). This indicates a reflection of natural usage of T cell receptor beta variable gene sequences in the patents.

We further investigated the TCR gene usage in the newly patented tebentafusp (18) by identifying its closest germline genes. The V-region genes of the alpha and beta chain have been identified as TRAV17-01 and TRBV19-01, respectively. These genes correspond to the third and the fourth mostly used V-region genes in published patents (Figure 2).

### Discussion and Conclusions

T-cell receptors are a novel class of biotherapeutic modalities that are gaining prominence with the recent approval of a first TCR therapeutic. Development of data resources for these molecules would certainly provide a sound basis for computational study of these molecules. In certain areas such resources already exist, for instance collating structures (9) or large-scale next-generation sequencing data (10). Specifically, collating therapeutic sequences can lead to insights regarding their therapeutic properties, as was the case with antibodies (19). Nevertheless, unlike with antibodies, of which there are more than 800 molecules with assigned INNs (20), there do not exist similar resources for TCR sequences. This is mostly due to the fact that there is only one approved therapeutic and merely a handful disclosed in development. Therefore, any resource providing any possible information on therapeutic TCRs should provide value to the community.

To bridge this gap, here we compiled a database of T-cell receptors from patents, which should hold information on molecules that are in-line to reach the clinical development stages in the future. Through doing so we revealed the broad trends in TCR patenting space, such as the increasing number of such patents in recent years, indicating a rise in interest in this biotherapeutic modality. The number of patent families with TCR sequence information at about 500 documents is still dwarfed by the number of antibody-related families extracted at the same time, which stood at more than 18,000 (11).

Nevertheless, our resource is not without its limitations in this sphere. Not all the TCRs that we identified held the classification as being used for therapeutic purposes. Furthermore, patented sequences are disclosed for legal protection rather than conveying engineering information, hindering attempts at pairing the sequences, or reliably identifying their targets. Despite such limitations, we consider our resource as a valuable addition to the computational TCR community as offering an early glimpse at molecules that might someday become therapeutic leads.

We hope that our compilation of patented TCR sequences will prove a valuable resource to facilitate the computational efforts to engineer these molecules for therapeutic use.

## Data Availability

The PATCRdb is freely available under the following github link: http://github.com/konradkrawczyk/patcrdb. The database is provided as .csv format listing the sequences and their metadata separately. Subsequent updates to the database will be pushed to this repository.

